# Hazardous Alcohol Use, Sexual Behavior, and Incident HIV across 11 Eastern and Southern African Countries

**DOI:** 10.64898/2026.03.30.26349734

**Authors:** Domonique M. Reed, Leigh F. Johnson, Katherine Keyes, Jesse Knight, Jeffrey W. Imai-Eaton

## Abstract

**Objectives:** Quantify hazardous alcohol consumption prevalence among individuals at risk of acquiring HIV infection and its association with high-risk sexual behaviors and incident HIV in 11 Eastern and Southern African countries.

**Design:** Secondary analysis of 16 nationally-representative household surveys (2015-2023).

**Methods:** The study included sexually active individuals aged ≥15 years. Alcohol use patterns were classified using the AUDIT-C (non-drinkers/low-risk drinkers/hazardous non-binge drinkers/hazardous binge drinkers). Outcomes included high-risk sexual behaviors, recent HIV infection, and undiagnosed HIV infection. Survey-weighted alcohol use prevalence and logistic regression were estimated by gender, adjusting for sociodemographic covariates. Model outputs were used to estimate change in incident infections when removing excess risks associated with alcohol use patterns.

**Results:** Analyses included 251,931 participants. Across countries, 5.8%-21.1% reported hazardous binge drinking, and 3.7%-15.7% reported hazardous non-binge drinking, with large gender differences. Sexual risk behaviors increased with drinking severity among men and women. Compared with non-drinkers, alcohol use was associated with higher odds of undiagnosed HIV infection; adjusted odds ratios ranged from 1.32 (1.16-1.50) for low-risk drinkers to 1.52 (1.34-1.72) for hazardous binge drinkers among men, and 1.28 (1.13-1.46) to 1.55 (1.31-1.82) among women. Simulated removal of alcohol-associated excess risk reduced undiagnosed HIV by 15.1% (10.9%-19.4%) among men and 5.8% (4.0%-7.9%) among women. Estimates for recent HIV infection followed a similar pattern but with larger uncertainty.

**Conclusions:** Hazardous alcohol use was associated with sexual risk and HIV infection in Eastern and Southern Africa. Reaching individuals who use alcohol with effective HIV prevention may reduce HIV acquisition risk across the region.

## Introduction

Alcohol consumption has been linked to HIV acquisition through its proximity to sex with new or non-regular partners and influence on decision-making about measures to prevent acquisition (e.g., condom use).[1–5] Alcohol use has been associated with heightened HIV acquisition risk globally, including across sub-Saharan Africa.[6–9] However, evidence was largely generated during earlier stages of the epidemic, is geographically and demographically fragmented, and is mostly concentrated in countries with established alcohol surveillance systems.[10,11] Studies also disproportionately focus on vulnerable populations or people living with HIV rather than the wider population at risk of acquiring HIV infection.[8,12–15] Rapid urbanization, expanding alcohol markets, and HIV prevention and treatment advances across the region motivate re-assessment of evidence on evolving consumption patterns, epidemic dynamics, and contemporary alcohol-HIV risk relationships.

Reported alcohol consumption varies widely across sub-Saharan Africa, with high rates of both abstention and episodic or heavy consumption.[14,16] These diverse patterns reflect variations in norms and practices, such as differences in alcohol regulation and policies, religious values and prohibitions, access to formal and informal alcohol markets, and prevailing gender norms that may stigmatize or restrict women’s public alcohol consumption while allowing or encouraging it among men.[17–20] This heterogeneity complicates defining relevant consumption metrics and characterizing epidemiologically relevant dimensions of use, limiting the development of effective interventions. Challenges include determining which dimensions of alcohol use severity (e.g., frequency, quantity, heavy episodic drinking) are most relevant to HIV acquisition risk and selecting validated assessment instruments.[15,21,22] Scarcity of comparable population-level data constrains efforts to characterize alcohol-related HIV risk pathways.[12,18]

Different drinking behaviors may confer varying risks for HIV acquisition.[23,24] Hazardous alcohol use refers to patterns of drinking that increase one’s risk of social, mental, or physical harm.[25,26] This construct encompasses two distinct but related forms of consumption: sustained heavy alcohol use reflects a chronic exposure pattern defined by regular alcohol intake exceeding low-risk drinking guidelines, while heavy episodic drinking, often termed as binge drinking, is characterized by acute consumption of large quantities of alcohol on a single occasion.[27] The chronic consumption patterns characterising sustained alcohol use may contribute to risk through long-term behavioral disinhibition, whereas acute, high-intensity binge drinking can precipitate immediate sexual risk-taking and exposure.[28–30] Addressing whether distinct hazardous patterns confer differential HIV risk through a gender-specific lens is essential, given gender differences in drinking patterns and sexual risk contexts.[6,31,32]

We analyzed data from 16 nationally-representative household surveys conducted between 2015 and 2023 across 11 Eastern and Southern African countries, the global region with the largest HIV burden. The three study objectives were to estimate the prevalence of hazardous alcohol consumption among adults at risk of acquiring HIV, assess associations between hazardous alcohol consumption and engagement in high-risk sexual behaviors, and examine associations between hazardous alcohol consumption and proxies of HIV incidence to assess whether observed behavioral risks translated into measurable infection risk.

## Methods

### Data

We selected surveys that had consistent measures of alcohol use, sexual risk behaviors, and HIV incidence, including the Botswana AIDS Indicator Survey (BAIS; 2021), South African HIV Behavioural, Sero-status and Media (SABSSM; 2017), and Population-based HIV Impact Assessment (PHIA) surveys in 10 countries between 2015 and 2023 (Figure 1). Survey details are published elsewhere.[33–48] These surveys were designed to measure the HIV epidemic and the impact of programs in high-burden countries. All surveys used two-stage stratified cluster sampling to obtain a representative sample of the national population. Consenting household members completed private face-to-face individual interviews and provided blood samples for HIV testing. HIV serologic testing was conducted at the time of survey, following country-specific national HIV testing guidelines. Blood samples collected from positive persons were tested in central labs for HIV viral load, the presence of antiretrovirals used in each country, and recent HIV infection status using the limiting antigen-avidity (LAg) enzyme immunoassay. HIV positive persons were classified as ‘recently infected’ if LAg normalized optical density value was ≤1.5, viral load was ≥1,000 copies/mL, and no serologic evidence of antiretroviral use.

**Figure 1.**
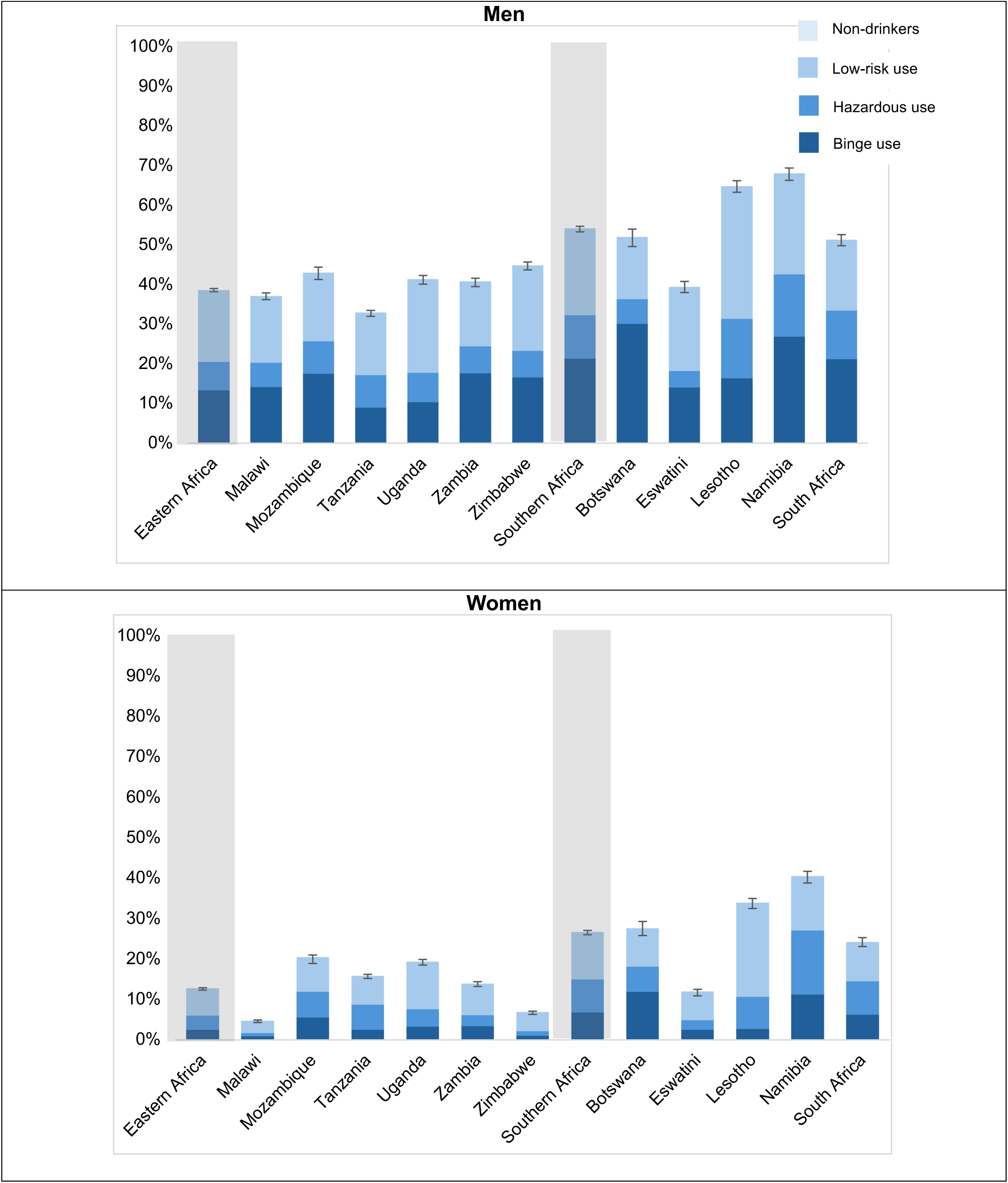
Survey-weighted Prevalence of Patterns of Hazardous Alcohol Use across by Gender and Country, N=245,253.

Our analysis included all ever sexually active individuals aged 15+ years, with varying upper age limits across surveys (see Supplemental Figure 1). Analyses were further restricted to participants who had complete information on HIV serostatus, alcohol use, and key demographic characteristics. Analyses estimating hazardous alcohol consumption prevalence and its associations with high-risk sexual behaviors were restricted to participants who tested HIV-negative in the survey (and were therefore at risk of acquiring HIV). Analyses examining patterns of HIV incidence additionally included individuals with evidence of lab-determined recent HIV infection or undiagnosed HIV infection.

### Measures

The primary exposure of interest was hazardous alcohol use, assessed with the Alcohol Use Disorder Identification-Consumption (AUDIT-C) scale.[49] AUDIT-C is based on three questions: 1) How often do you have a drink containing alcohol? (Never/Monthly or less/2-4 times a month/2-3 times a week/4 or more times a week); 2) How many drinks containing alcohol do you have on a typical day? (1 or 2/3 or 4/5 or 6/7 to 9/10 or more); and 3) How often do you have (for men) five or more and (for women) four or more drinks on one occasion? (Never/Less than monthly/Monthly/Weekly/Daily or almost daily). Item responses were summed to classify alcohol use as non-drinking (score = 0), low-risk drinking (score <4 for men or <3 for women), or hazardous drinking (score ≥4 for men or ≥3 for women). Hazardous drinkers were further categorized based on the third item, identifying individuals who reported heavy episodic drinking at least monthly as ‘hazardous binge drinkers’. We combined the classifications into four categories: *non-drinkers, low-risk drinkers, hazardous non-binge drinkers, and hazardous binge drinkers*.

We considered two outcomes of interest: engaging in high-risk sexual behaviors and incident HIV. Sexual risk behavior was assessed through four binary self-reported outcomes: two or more sexual partners in the past year; sold or bought sex in the past year; condomless sex with a non-marital or non-cohabitating partner in the past year; and alcohol used by self or partner at last sex. Specific survey questions used are in the Supplementary Table S1. Botswana, Mozambique, and Uganda surveys did not capture whether individuals sold or bought sex in the past year, and the Namibia survey did not capture whether alcohol was used at last sex. Consequently, the analytic samples vary slightly across outcomes.

Incident HIV was assessed using two proxy indicators: recent HIV infection status and undiagnosed HIV infection. Biomarker assays for recent infection detect new infections within roughly four months, offering high specificity but yielding few cases, limiting power for association analysis. Conversely, using undiagnosed HIV infection increases the number of positive outcomes and the ability to identify associations, but may include some individuals who acquired HIV years ago but had not yet been diagnosed (or did not disclose their diagnosis). For undiagnosed HIV, individuals were classified as undiagnosed if they tested positive for HIV but reported to be unaware of their status and did not have antiretrovirals detected. South Africa was excluded from the recent infection analysis because the publicly available SABSSM dataset did not include recent infection status. Within SABSSM, respondents were classified as undiagnosed if they tested HIV-positive and reported never having been tested, receiving a negative result at their last test, or not receiving their most recent test results.

Adjusted models included variables commonly associated with our exposures and outcomes of interest: age (15-24 years/25-34 years/35-49 years/50+ years), education completed (None or primary/Secondary/More than secondary), employment status (Unemployed or unable to work/Employed or student), residence type (Urban/Rural), age at first sex (≤15 years/>15 years), and country. Sexual risk behaviors were not adjusted for in the incident HIV models, as those variables are likely mediators between alcohol use and acquisition.

### Statistical Analysis

All analyses were stratified by gender, as reported in the survey, reflecting systematic differences in alcohol consumption and transmission patterns. Analyses accounted for complex survey design using the *survey* package in R (version 4.12.0).[50] To pool multiple surveys, we normalized survey weights to the total sample size in each survey, ensuring that each survey contributed comparably to the analyses. Gender-stratified prevalence of alcohol use, sociodemographic characteristics, and self-reported engagement in sexual behaviors were summarized by non-weighted frequency and survey-weighted column percentages and design-based 95% confidence intervals (CIs). Model analyses used survey-weighted logistic regression to estimate unadjusted and adjusted odds ratios (ORs) and 95% CIs. Additionally, we used model outputs to simulate proportionally how much lower recent and undiagnosed infections would be if excess risks associated with various alcohol use patterns were set to the level of non-drinkers, with uncertainty estimated via Monte Carlo simulation of the adjusted regression coefficients. We report both level-specific estimates and an overall estimate representing the joint effect across alcohol use categories. Statistical significance was determined by whether the 95% confidence interval excluded the null value.

The Harvard Longwood IRB determined this analysis of secondary data as non-human subjects research (IRB Protocol #: IRB26-0109).

## Results

### Sample Characteristics

Data were available from 358,894 adults aged 15+ years across 16 population-based surveys. Of those, 245,253 were HIV-negative, had ever had sex, and had complete information on relevant outcomes and covariates. The survey-weighted proportion of men was 48.2% (Table 1; unweighted n=104,200), and 51.8% were women (unweighted n=141,053). Almost 30% were aged 25-34 years. Most participants had no or only primary education (51.4%; men: 47.6%, women: 55.0%), were unemployed or unable to work (55.2%; men: 44.0%, women: 65.6%), lived in rural areas (61.0%; men: 61.1%, women: 60.9%), had ever been married (69.6%; men: 62.4%, women: 76.4%), and resided in East African countries (71.1%; men: 70.8%, women: 71.4%).

**Table 1.**
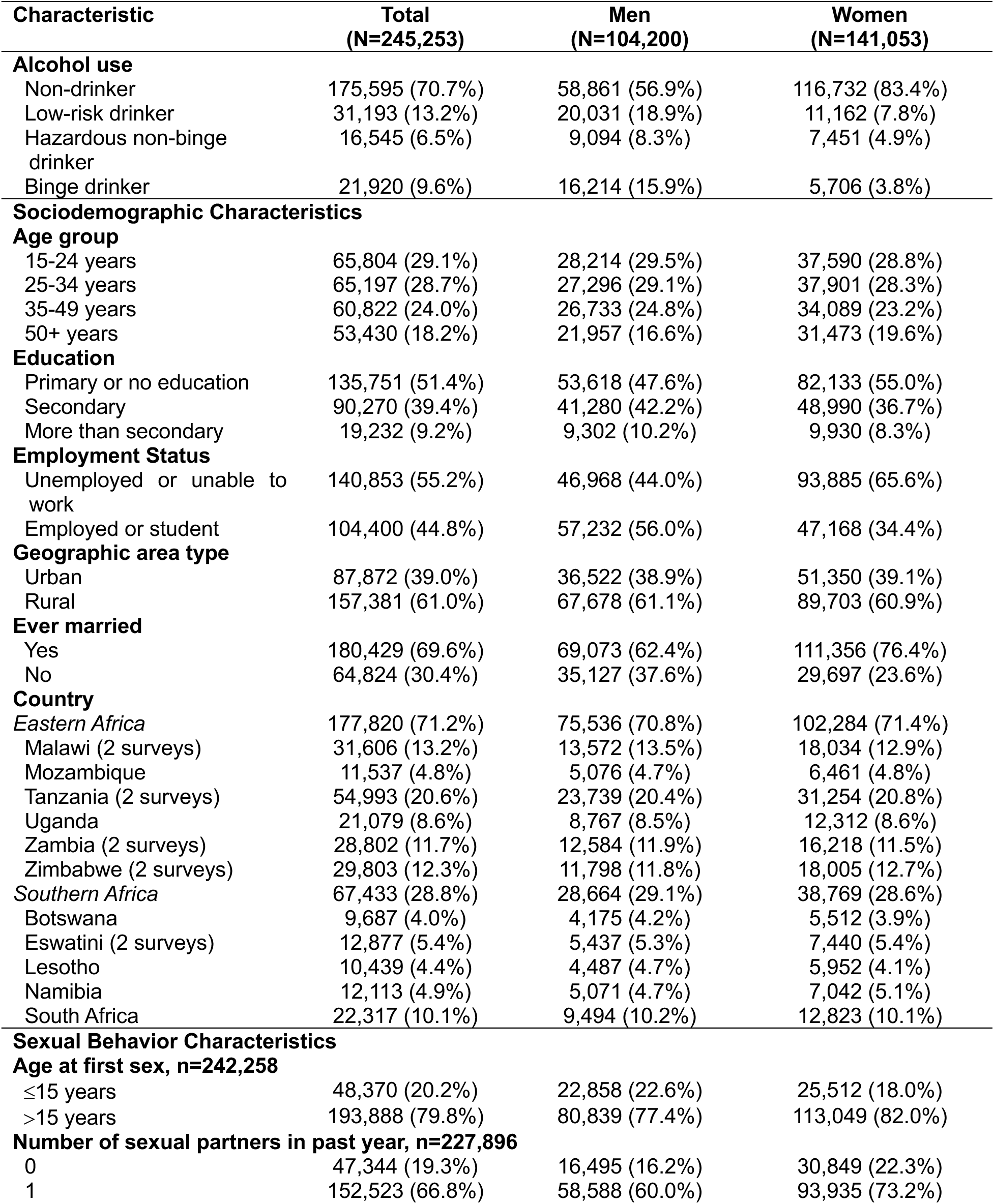
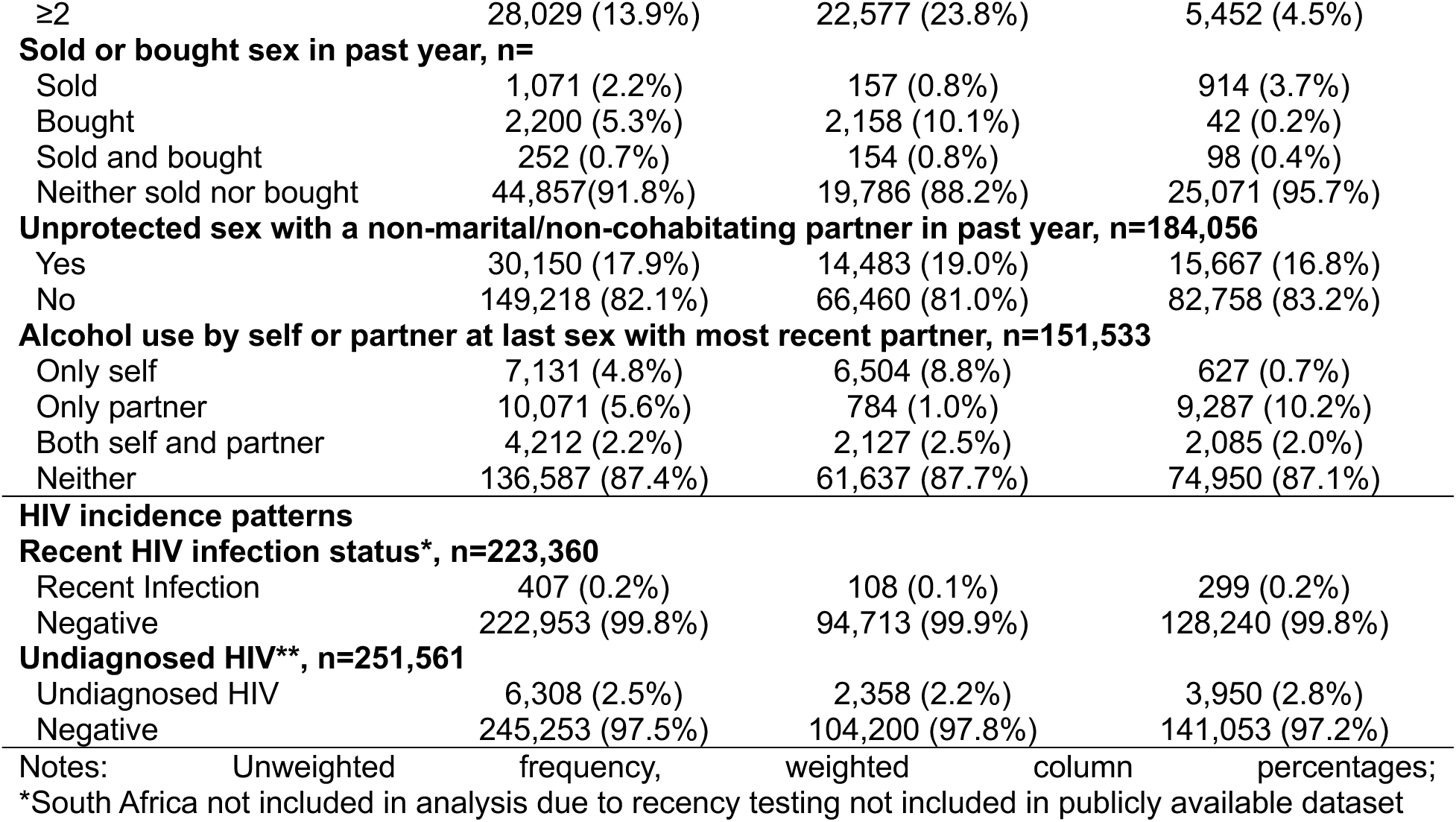
Characteristics of HIV-negative survey participants aged 15 years or older who have ever had sex across 16 surveys conducted in 11 Eastern and South African countries, by gender, N=245,253.

### Prevalence of Hazardous Alcohol Use

Most sexually-active individuals reported being non-drinkers (70.7%; men: 56.9%, women: 83.4%; Table 1), followed by low-risk drinkers (13.2%; men: 18.9%, women: 7.8%), hazardous binge drinkers (9.6%; men: 15.9%, women: 3.8%), and hazardous non-binge drinkers (6.5%; men: 8.3%, women: 4.9%). A higher proportion were non-drinkers in Eastern Africa than in Southern Africa (75.0% versus 60.1%; Table 2). A greater proportion in Southern Africa reported hazardous binge drinking (14.0% versus 7.8%), hazardous non-binge drinking (9.5% versus 5.4%), and low-risk drinking (16.8% versus 11.8%).

**Table 2.**
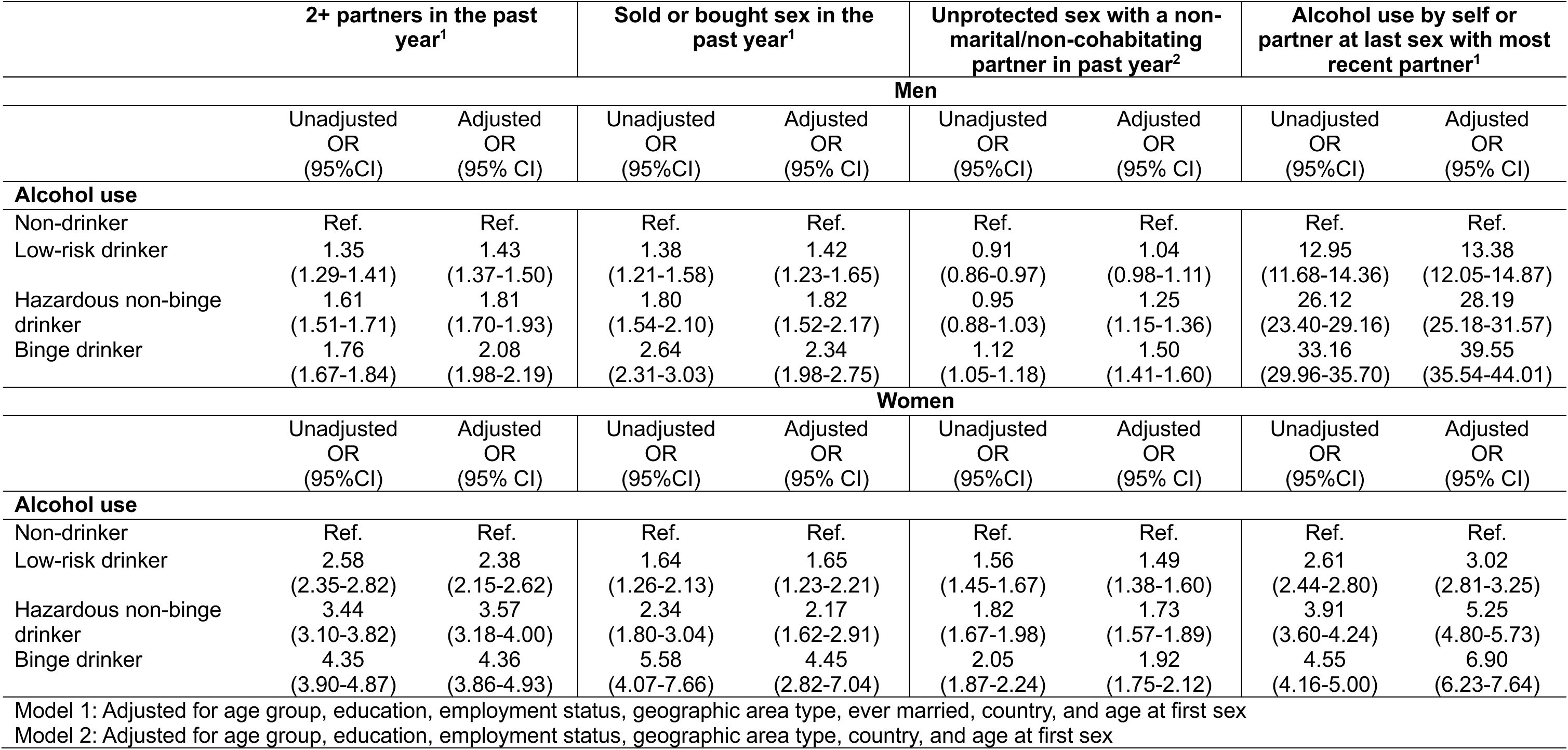
Association Between Patterns of Hazardous Alcohol Use and Risky Sexual Behaviors among People who are HIV Negative by Gender, n=245,253.

Across all countries, men were more likely than women to report all drinking patterns; however, prevalence varied widely across countries. Low-risk drinking among men ranged from 15.5% in Botswana to 33.1% in Lesotho (Figure 1), and, among women, from 2.7% in Malawi to 23.2% in Lesotho. Hazardous non-binge drinking among men ranged from 4.3% in Eswatini to 15.9% in Namibia, and, among women, from 0.8% in Malawi to 15.7% in Namibia. Prevalence of hazardous binge drinking among men ranged from 9.1% in Tanzania to 27.1% in Botswana, and among women, from 1.1% in Malawi to 11.7% in Namibia.

### Sexual Risk Behaviors

Among ever sexually active individuals at risk of acquiring HIV, 13.9% (men: 23.8%, women: 4.5%; Table 1) reported two or more partners in the past year, 8.2% (men: 11.7%, women: 4.3%) sold or bought sex in the past year, 17.9% (men: 19.0%, women: 16.8%) had condomless sex with a non-marital or non-cohabitating partner at last sex, and 12.6% (men: 12.3%, women: 12.9%) reported alcohol use by self or partner at last sex.

More severe alcohol use was associated with higher odds of all sexual risk behaviors among both genders, adjusted for other covariates (Table 3). Among men, low-risk drinkers, hazardous non-binge drinkers, and hazardous binge drinkers had 1.43 (adjusted 95% CI: 1.37-1.50), 1.81 (1.70-1.93), and 2.08 (1.98-2.18), respectively, higher odds of reporting two or more partners in the past year compared to non-drinkers. Low-risk drinkers, hazardous non-binge drinkers, and hazardous binge drinkers had 1.42 (1.23-1.65), 1.82 (1.52-2.17), and 2.34 (1.98-2.19), respectively, higher odds of selling or buying sex in the past year. Hazardous non-binge drinkers and hazardous binge drinkers had 1.25 (1.15-1.36) and 1.50 (1.41-1.60), respectively, higher odds of having condomless sex with a non-marital or non-cohabitating partner at their most recent sex compared to non-drinkers. However, there was no or minimal difference in reporting high-risk sex among low-risk drinkers compared to non-drinkers (OR: 1.04; 95% CI: 0.98-1.11). Low-risk drinkers, hazardous non-binge drinkers, and hazardous binge drinkers had 13.38 (12.05-14.87), 28.19 (25.18-31.57), and 39.55 (35.54-44.01), respectively, higher odds of alcohol use by self or partner at last sex with most recent partner.

**Table 3.**
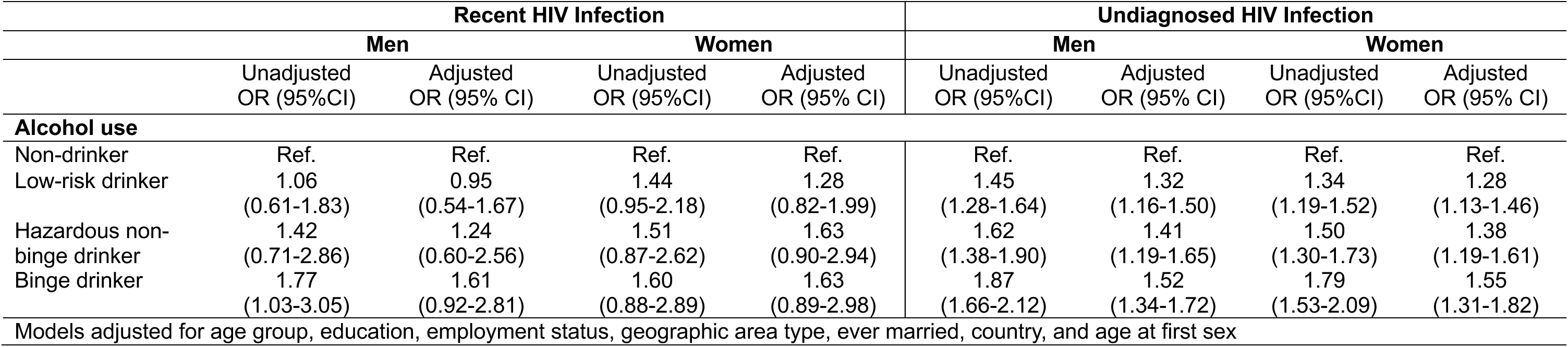
Association Between Patterns of Hazardous Alcohol Use and Recent and Undiagnosed HIV Infection by Gender, n=251,931.

While alcohol use prevalence was lower among women, the magnitudes of associations with sexual risk were generally larger (Table 3). Low-risk drinkers, hazardous non-binge drinkers, and hazardous binge drinkers had 2.38 (2.15-2.62), 3.57 (3.18-4.00), and 4.36 (3.86-4.93), respectively, higher odds of two or more partners in the past year compared to non-drinkers. They had 1.65 (1.23-2.21), 2.17 (1.62-2.91), and 4.45 (2.82-7.04), respectively, higher odds of selling or buying sex in the past year compared to non-drinkers, and had 1.49 (1.38-1.60), 1.73 (1.57-1.89), and 1.92 (1.75-2.12), respectively, higher odds of condomless sex with a non-marital or non-cohabitating partner. Low-risk drinkers, hazardous non-binge drinkers, and hazardous binge drinkers had 3.02 (2.81-3.25), 5.25 (4.80-5.73), and 6.90 (6.23-7.64), respectively, higher odds of alcohol use by self or partner at last sex with most recent partner compared to non-drinkers.

### Patterns of HIV Incidence

Across the surveys, 0.2% (n=407; men: 0.1%, women: 0.2%; Table 1) of individuals were classified as having a recent HIV infection, and 2.5% (n=6,308; men: 2.2%, women: 2.8%) had undiagnosed HIV infection. Among men with recent infection, 50.0% were non-drinkers, while 22.4% were hazardous binge drinkers, 9.8% hazardous non-binge drinkers, and 17.8% low-risk drinkers; among women, the large majority (78.6%) were non-drinkers, with much smaller proportions reporting hazardous binge (5.1%), hazardous non-binge (6.4%), or low-risk drinking (9.9%). The pattern was similar for undiagnosed HIV infection; among men 23.0% with undiagnosed HIV engaged in hazardous binge 10.8% hazardous non-binge, 21.5% low-risk, and 44.7% non-drinkers compared to (77.1%) of women with undiagnosed HIV were non-drinkers, and relatively few reported hazardous binge (6.2%), hazardous non-binge (6.9%), or low-risk drinking (9.8%).

Among men, hazardous non-binge drinkers and hazardous binge drinkers had 1.24 (0.60-2.53; Table 4) and 1.61 (0.92-2.81) higher adjusted odds of recent HIV infection compared to non-drinkers; however, low-risk drinkers were not associated with recent HIV infection (OR: 0.95; 95% CI: 0.54-1.68). Among men, removing the excess risk associated with alcohol use reduced recent infections by 9.1% (−9.4-28.8%; Figure 2 shows use pattern-specific estimates). Among women, low-risk drinkers, hazardous non-binge drinkers, and hazardous binge drinkers had 1.28 (0.82-1.99), 1.63 (0.90-2.94), and 1.63 (0.89-2.98) higher odds of recent HIV infection compared to non-drinkers. Removing alcohol-associated excess risk reduced recent infections by 6.7% (1.0-14.7%).

**Figure 2.**
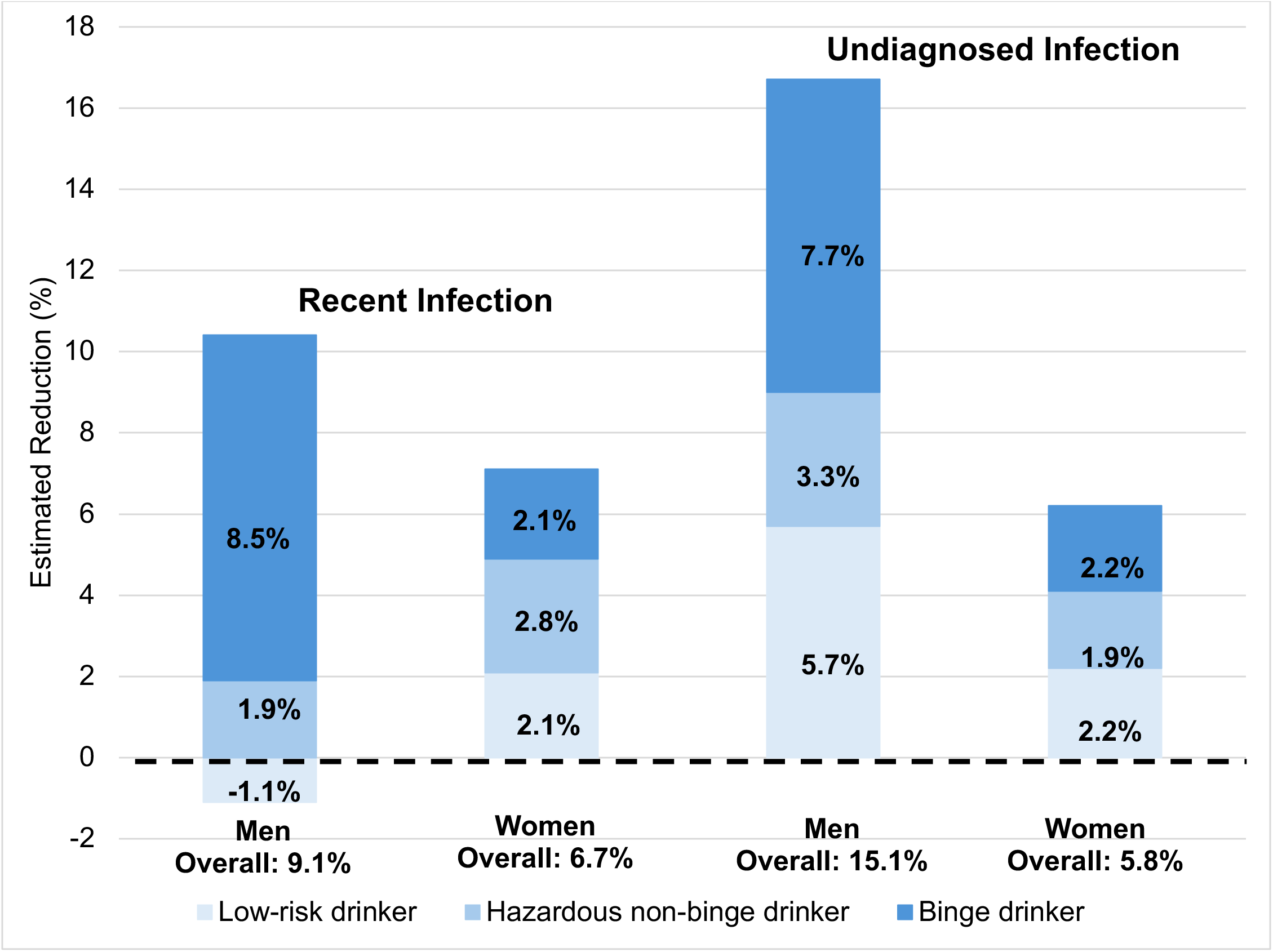
Estimated Reduction (%) in Recent and Undiagnosed HIV Infections if Excess Risk Associated with Alcohol Use Patterns Were Removed, by Gender across 11 Eastern and Southern African Countries (2015–2023), N = 251,931.

Statistical relationships were similar but more precise for undiagnosed HIV infection due to larger numbers. Among men, low-risk drinking, hazardous non-binge drinking, and hazardous binge drinking had 1.32 (1.16-1.50), 1.41 (1.19-1.65), and 1.52 (1.34-1.72) higher odds compared to non-drinkers. Removing alcohol use-associated excess risk reduced undiagnosed infection among men by 15.1% (10.9-19.4%). Among women, low-risk drinking, hazardous non-binge drinking, and hazardous binge drinking had 1.28 (1.13-1.46), 1.38 (1.19-1.61), and 1.55 (.31-1.82) higher odds of undiagnosed HIV infection compared to non-drinkers. Removing alcohol-associated excess risk reduced the undiagnosed HIV infection among women by 5.8% (4.0-7.9%).

## Discussion

Alcohol consumption varied across countries in Eastern and Southern Africa, with higher consumption in Southern Africa, and consistently higher consumption among men than women. Among both genders, alcohol use severity was associated with progressively greater engagement in behaviors associated with HIV acquisition, despite marked differences in alcohol use. A similar dose-response relationship was observed between alcohol use patterns and undiagnosed HIV infection. Estimates for recent HIV infection followed a similar pattern, but confidence intervals were wide and did not provide statistical evidence of a population-level association.

Few studies have estimated hazardous alcohol use prevalence across multiple Eastern and Southern African countries.[51,52] The pronounced consumption difference between men and women is consistent with prior studies in this region.[14,53] Most adults reported being non-drinkers (57% of men; 83% of women), exceeding the World Health Organization’s global estimates that 48% of adult men and 65% of women abstained from alcohol in 2019.[54] The magnitude of the gender gap varied across countries, with the narrowest in Tanzania and the widest in Zimbabwe. While Tanzania’s narrow gap reflects high non-drinking among both genders, other variations in gender gaps may reflect country-specific norms shaping drinking behavior. In Zimbabwe, for example, alcohol is consumed socially in male-dominated venues like beer halls.[55] These entrenched norms and contexts may also drive women to underreport their alcohol use overall or minimize the severity of use, potentially obscuring actual consumption patterns. In other global regions, evidence is growing of a narrowing alcohol use gender gap;[14,56–58] future research should assess whether similar trends arise across this region of Africa, and, if so, the implications for HIV risk dynamics identified here.

Hazardous alcohol use among men, including sustained heavy drinking and binge drinking, was high, ranging from 17% in Tanzania to 43% in Namibia. Men more often engaged in binge drinking than sustained heavy drinking alone. In contrast, women who report hazardous alcohol use showed more varied patterns across countries. The concentration of men’s hazardous binge drinking may arise from social norms that promote heavy episodic drinking in group settings and peer contexts where such behavior is culturally acceptable. Previous research has associated psychological stressors and experiences of violence with hazardous alcohol use among both women and men, although the nature and social acceptability of drinking in response to these stressors may differ by gender and setting.[59–61] These different and potentially overlapping pathways have implications for HIV prevention efforts. Effective strategies may need to address psychosocial stressors and trauma (e.g., through screening and referral to mental health and violence-related services) alongside the social contexts where drinking takes place, including public, peer-oriented venues where drinking might be more socially acceptable among men, like bars or drinking establishments, as well as less visible or private drinking settings that could be more common among women, such as drinking in household or small social environments.

A clear dose-response emerged between increasingly hazardous alcohol consumption and reporting multiple sexual partners, transactional sex, unprotected sex with non-cohabiting partners, alcohol use during sex, and incident HIV proxies; and the strength of associations was even stronger among women than among men. Given both the elevated risk and the large proportion engaging in binge drinking, particularly among men, interventions that mitigate HIV acquisition risk among this group could potentially avert a meaningful share of new infections.[23,28] Gender differences in reporting of which partner used alcohol during sex may reflect the intersection of alcohol use with other vulnerabilities, including gender-based power imbalances, economic instability, and experiences of violence, and gendered patterns in drinking and sexual encounters.[23,62]

All forms of alcohol use were associated with higher odds of recent infection and undiagnosed HIV infection. The association with undiagnosed HIV infection may arise through both higher incidence and lower testing uptake and delayed diagnosis among people who drink alcohol.[53] Together, these findings highlight the importance of ensuring both prevention, HIV testing, and other care services effectively reach people who drink.[63] Innovative approaches may be needed for reaching individuals across the full spectrum of alcohol use severity who are currently being missed by existing strategies.

Our counterfactual analysis quantified the share of infections associated with excess incidence risk among alcohol users to inform the potential epidemic-level impact of reaching this group with effective HIV prevention. Causally attributing excess HIV acquisition and transmission risk to alcohol use—and hence potential epidemiologic impact of effective interventions reducing alcohol use—is more uncertain and difficult to assess given underlying factors that shape alcohol use, sexual risk, and HIV acquisition, which were not measured in our data. These factors include norms around masculinity, individual risk propensity, and religion. For example, endorsing inequitable gender norms and personality factors have been shown to be associated with both increased binge drinking and sexual risk behaviour.[19,20,64–68] Moreover, reverse causality is plausible, with individuals frequenting drinking venues to meet sexual partners or drinking heavily as a psychosocial response to engaging in risky sex.[5,23]

Several further limitations warrant consideration when interpreting these findings. First, alcohol use and sexual behaviors were assessed through self-report and may be subject to recall error and social desirability bias.[69,70] Self-reported alcohol consumption, including measures based on the AUDIT-C, may underestimate both drinking frequency and intensity.[71] Second, gender identities were not captured in most surveys, limiting our ability to examine alcohol use and HIV risk among gender minority populations. Although the SABSSM survey permitted reporting of transgender and intersex identities, the small number of respondents selecting these categories (n = 37) precluded analysis of these gender identities. Third, although we harmonized variables across surveys to enable cross-country comparisons, variation in some survey items may have introduced heterogeneity in our estimates. Fourth, some participants did not respond to the questions on sexual risk behavior, potentially introducing selection bias. Moreover, non-response could be differential by gender and country context, depending on local norms.

## Conclusion

Hazardous alcohol use was prevalent across Eastern and Southern Africa, and its severity was associated with sexual risk and proxies of incident HIV. While associations were often stronger among women, higher alcohol use among men translated into larger proportions of infections from alcohol-associated excess risks. These findings point to the need for gender-specific interventions that integrate HIV prevention and testing into alcohol reduction programming and deliver services in communities and social settings where drinking occurs.

## Acknowledgements

D.M.R., J.W.I-E., and L.F.J conceptualized the study. D.M.R conducted the analyses and drafted the manuscript. L.F.J., K.K., J.K., and J.W.I-E contributed to the interpretation of the findings and critically revised the manuscript. All authors reviewed and approved the final version of the manuscript.

## Data availability

The survey datasets analyzed in this study are publicly available from the respective survey programs upon request or registration.

