## Supplemental Material for "Hazardous Alcohol Use, Sexual Behavior, and Incident HIV across 11 Eastern and Southern African Countries"

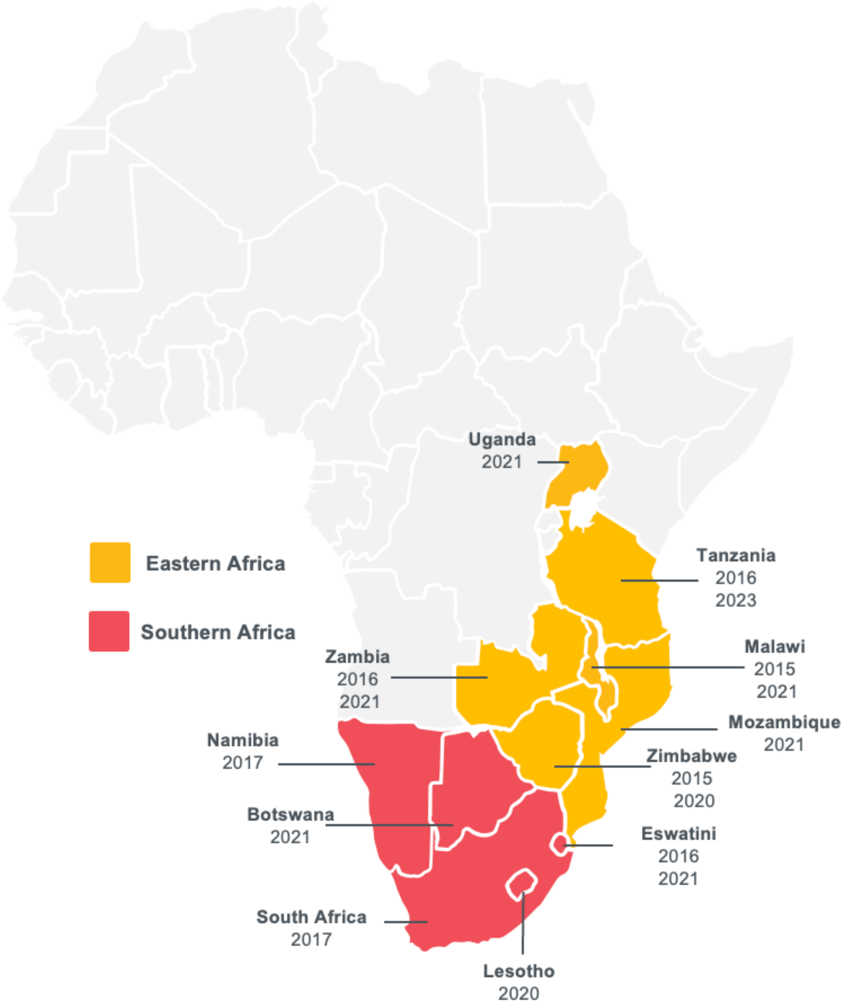

**Figure S1.** Eastern and South African countries included in the study and years in which surveys were conducted. Data were included from the South African HIV Behavioral, Sero-status and Media Survey (SABSSM; Wave V), the Botswana AIDS Impact Survey (BAIS; Wave V), and the Population-based HIV Impact Assessment surveys (PHIA; Wave 1 and 2). The adult study population included individuals aged 15 years or older, with varying upper age limits across surveys. The Zambia (2016) survey enrolled individuals up to 59 years; the Botswana (2021), Malawi (2015), and Namibia (2017) surveys enrolled individuals up to 64 years, and all other surveys had no upper age limit for study inclusion.

**Supplemental Material Section 1**. Operationalization of Sexual Risk Behavior Questions

Sexual risk behavior was assessed through four binary outcomes: having two or more sexual partners in the past year, sold or bought sex in the last 12 months; unprotected sex with a non-marital or non-cohabitating partner in the past year; and alcohol use by self or most recent partner at last sex. For sold or bought sex in the last 12 months, participants were asked separately if they had sold (or received money, gifts, or favours in exchange for) sex in the past year, and if they had bought (or given or received money, gifts, or favours in exchange for) sex. If participants answered affirmatively to either of these questions, then they were categorized as Yes for sold or bought sex in the last 12 months. For unprotected sex with a non-marital or non-cohabitating partner, participants were asked about their most recent sexual partner and were asked to characterize the relationship with this partner (e.g., husband or wife, live-in partner, sex worker, friend), and then relationship types were categorized as non-marital or non-cohabitating. Then participants were asked with that same partner, if a condom was used during the most recent sex with that partner. Using these two questions, we categorized individuals into a high-risk sexual behavior of unprotected sex with a non-marital or non-cohabitating partner. For alcohol use by self or most recent partner at last sex, participants were asked if at the last time they had sex with their most recent partner, did either of you drink alcohol beforehand, and the participants could respond as only they were drinking, only their partner was drinking, both of them were drinking, or neither. Alcohol use by self, partner, or both was categorized as alcohol use at the last sex with their most recent partner.

**Table S1.** Survey-specific Questions for Sexual Behavior and HIV Incidence Outcomes

| | **Outcomes** | | | | | |
|---|---|---|---|---|---|---|
| **Survey** | Number of sexual partners in the past year | Ever sold sex or buy sex | Unprotected sex with a non-marital/non-cohabitating partners in the past year | Alcohol use at last sex with most recent partner | Recent HIV infection | Undiagnosed HIV |
| BAIS | How many different people have you had sex with in the last 12 months? | - | The last time you had sex with [initials], was a condom used? (Yes; No)<br><br>What is your relationship with your [initials] partner? (Husband/Wife; Live-in partner; Partner, not live-in; Ex-spouse/Ex-partner; Friend/ Acquaintance; Stranger; Other) | The last time you had sex with [initials], did either of you use any drugs, alcohol, or substances beforehand? (Only I was; Only partner was; Both were; Neither) | LAg (Positive: recent; Positive: long-term) | HIV-positive and unaware and antiretroviral not detectable or antiretroviral testing results missing |
| SABSSM VI | Have you had sex during the past 12 months? (Yes; No)<br><br>Overall, how many sexual partners did you have during the past 12 months? | Have you ever received money, gifts, or favours in exchange for sex? (Yes; No)<br><br>In the last 12 months, have you received money, gifts, or favours in exchange for sex? (Yes; No)<br><br>Have you ever given money, | Did you use a condom at last sex? Most recent (Yes; No)<br><br>Can you describe this partner? Most recent person with whom you had sex (Husband/Wife; Live-in partner; Girlfriend/Boyfriend not living with you; Casual partner; Someone whom | The last time you had sex with your partner did you drink alcohol before sex? Most recent (Yes; No) | - | HIV-positive, and never had a previous HIV test, previously tested negative for HIV, or never received HIV result |

| | | | | | | |
|---|---|---|---|---|---|---|
| | | gifts, or favours in exchange for sex? (Yes; No)<br><br>In the last 12 months, have you given money, gifts, or favours in exchange for sex? (Yes; No) | you paid for sex; Other) | | | |
| PHIA | | | | | | |
| Eswatini 2016 | In total, with how many different people have you had sex with in the last 12 months? | Have you ever sold sex for money? (Yes; No)<br><br>In the last 12 months, have you sold sex for money? (Yes; No)<br><br>Have you ever paid money for sex? (Yes; No)<br><br>In the last 12 months, have you paid money for sex? (Yes; No) | The last time you had sex with [initials], was a condom used? (Yes; No)<br><br>What is your relationship with your [initials] partner? (Husband/Wife; Live-in partner; Partner, not live-in; Ex-spouse/Partner; Friend/ Acquaintance; Sex worker; Sex worker client; Stranger; Other) | - | LAg (Positive: recent; Positive: long-term) | HIV-positive and unaware and antiretroviral not detectable or antiretroviral testing results missing |
| Eswatini 2021 | How many different people have you had sex with in the last 12 months? | - | The last time you had sex with [initials], was a condom used? (Yes; No)<br><br>What is your relationship with your [initials] | The last time you had sex with [initials], did either of you drink alcohol beforehand? (Only I was drinking; Only partner was | LAg (Positive: recent; Positive: long-term) | HIV-positive and unaware and antiretroviral not detectable or antiretroviral testing results missing |

| | | | | | | |
|---|---|---|---|---|---|---|
| | | | partner? (Husband/Wife; Live-in partner; Partner, not live-in; Ex-spouse/Ex-partner; Friend/ Acquaintance; Sex worker; Sex worker client; Stranger; Other) | drinking; Both were drinking; Neither) | | |
| Lesotho 2020 | How many different people have you had sex with in the last 12 months? | Have you ever had sex for money and/or gifts or favours? (Yes;No)<br><br>In the last 12 months, have you had sex for money and/or gifts or favours? (Yes; No)<br><br>Have you ever paid money or given gifts/favours for sex? (Yes; No)<br>In the last 12 months, have you paid money or given gifts or favours for sex? (Yes; No) | The last time you had sex with [initials], was a condom used? (Yes; No)<br><br>What is your relationship with your [initials] partner? (Husband/Wife; Live-in partner; Partner, not live-in; Ex-spouse/Ex-partner; Friend/ Acquaintance; Sex worker; Sex worker client; Stranger; Other) | The last time you had sex with [initials], did either of you drink alcohol beforehand? (Only I was drinking; Only partner was drinking; Both were drinking; Neither) | LAg (Positive: recent; Positive: long-term) | HIV-positive and unaware and antiretroviral not detectable or antiretroviral testing results missing |
| Malawi 2015 | In total, with how many different people have you had sex with in the last 12 months? | Have you ever sold sex for money? (Yes; No)<br><br>In the last 12 months, have you | The last time you had sex with [initials], was a condom used? (Yes; No) | The last time you had sex with [initials], did either of you drink alcohol beforehand? (Only I was | LAg (Positive: recent; Positive: long-term) | HIV-positive and unaware and antiretroviral not detectable or antiretroviral testing results missing |

| | | | | | | |
|---|---|---|---|---|---|---|
| | | sold sex for money? (Yes; No)<br><br>Have you ever paid money for sex? (Yes; No)<br><br>In the last 12 months, have you paid money for sex? (Yes; No) | What is your relationship with your [initials] partner? (Husband/Wife; Live-in partner; Partner, not living with respondent; Ex-spouse/Partner; Friend/ Acquaintance; Sex worker; Sex worker client; Stranger; Other) | drinking; Only partner was drinking; Both were drinking; Neither) | | |
| Malawi 2021 | How many different people have you had sex with in the last 12 months? | - | The last time you had sex with [initials], was a condom used? (Yes; No)<br><br>What is your relationship with your [initials] partner? (Husband/Wife; Live-in partner; Partner, not live-in; Ex-spouse/Ex-partner; Friend/ Acquaintance; Sex worker; Sex worker client; Stranger; Other) | The last time you had sex with [initials], did either of you drink alcohol beforehand? (Only I was drinking; Only partner was drinking; Both were drinking; Neither) | LAg (Positive: recent; Positive: long-term) | HIV-positive and unaware and antiretroviral not detectable or antiretroviral testing results missing |
| Mozambique 2021 | How many different people have you had sex with in the last 12 months? | - | The last time you had sex with [initials], was a condom used? (Yes; No) | The last time you had sex with [initials], did either of you drink alcohol beforehand? | LAg (Positive: recent; Positive: long-term) | HIV-positive and unaware and antiretroviral not detectable or antiretroviral |

| | | | | | | |
|---|---|---|---|---|---|---|
| | | | What is your relationship with your [initials] partner? (Husband/Wife; Live-in partner; Partner, not live-in; Ex-spouse/Ex-partner; Friend/ Acquaintance; Sex worker; Sex worker client; Stranger; Other) | (Only I was drinking; Only partner was drinking; Both were drinking; Neither) | | testing results missing |
| Namibia 2017 | In total, with how many different people have you had sex with in the last 12 months? | Have you ever sold sex for money? (Yes; No)<br><br>In the last 12 months, have you sold sex for money? (Yes; No)<br><br>Have you ever paid money for sex? (Yes; No)<br><br>In the last 12 months, have you paid money for sex? (Yes; No) | The last time you had sex with [initials], was a condom used? (Yes; No)<br><br>What is your relationship with your [initials] partner? (Husband/Wife; Live-in partner; Partner, not live-in; Ex-spouse/Ex-partner; Friend/ Acquaintance; Sex worker; Sex worker client; Stranger; Other) | - | LAg (Positive: recent; Positive: long-term) | HIV-positive and unaware and antiretroviral not detectable or antiretroviral testing results missing |
| Tanzania 2016 | In total, with how many different people have you had sex with in the last 12 months? | In the last 12 months, have you sold sex for money? (Yes; No)<br><br>In the last 12 months, have you | The last time you had sex with [initials], was a condom used? (Yes; No)<br><br>What is your relationship with | The last time you had sex with [initials], did either of you drink alcohol beforehand? (Only I was drinking; Only | LAg (Positive: recent; Positive: long-term) | HIV-positive and unaware and antiretroviral not detectable or antiretroviral testing results missing |

| | | | | | | |
|---|---|---|---|---|---|---|
| | | paid money for sex? (Yes; No) | your [initials] partner? (Husband/Wife; Live-in partner; Partner, not live-in; Ex-spouse/Ex-partner; Friend/ Acquaintance; Sex worker; Sex worker client; Stranger; Other) | partner was drinking; Both were drinking; Neither) | | |
| Tanzania 2023 | How many different people have you had sex with in the last 12 months? | - | The last time you had sex with [initials], was a condom used? (Yes; No)<br><br>What is your relationship with your [initials] partner? (Husband/Wife; Live-in partner; Partner, not live-in; Ex-spouse/Ex-partner; Friend/ Acquaintance; Sex worker; Sex worker client; Stranger; Other) | The last time you had sex with [initials], did either of you drink alcohol beforehand? (Only I was drinking; Only partner was drinking; Both were drinking; Neither) | LAg (Positive: recent; Positive: long-term) | HIV-positive and unaware and antiretroviral not detectable or antiretroviral testing results missing |
| Uganda 2021 | How many different people have you had sex with in the last 12 months? | - | The last time you had sex with [initials], was a condom used? (Yes; No)<br><br>What is your relationship with your [initials] partner? | The last time you had sex with [initials], did either of you drink alcohol beforehand? (Only I was drinking; Only partner was drinking; Both | LAg (Positive: recent; Positive: long-term) | HIV-positive and unaware and antiretroviral not detectable or antiretroviral testing results missing |

| | | | | | | |
|---|---|---|---|---|---|---|
| | | | (Husband/Wife; Live-in partner; Partner, not live-in; Ex-spouse/Ex-partner; Friend/ Acquaintance; Sex worker; Sex worker client; Stranger; Other) | were drinking; Neither) | | |
| Zambia 2016 | In total, with how many different people have you had sex with in the last 12 months? | Have you ever sold sex for money? (Yes; No)<br><br>In the last 12 months, have you sold sex for money? (Yes; No)<br><br>Have you ever paid money for sex? (Yes; No)<br><br>In the last 12 months, have you paid money for sex? (Yes; No) | The last time you had sex with [initials], was a condom used? (Yes; No)<br><br>What is your relationship with your [initials] partner? (Husband/Wife; Live-in partner; Partner, not live-in; Ex-spouse/Ex-partner; Friend/ Acquaintance; Sex worker; Sex worker client; Stranger; Other) | The last time you had sex with [initials], did either of you drink alcohol beforehand? (Only I was drinking; Only partner was drinking; Both were drinking; Neither) | LAg (Positive: recent; Positive: long-term) | HIV-positive and unaware and antiretroviral not detectable or antiretroviral testing results missing |
| Zambia 2021 | How many different people have you had sex with in the last 12 months? | - | The last time you had sex with [initials], was a condom used? (Yes; No)<br><br>What is your relationship with your [initials] partner? (Husband/Wife; Live-in partner; | The last time you had sex with [initials], did either of you drink alcohol beforehand? (Only I was drinking; Only partner was drinking; Both were drinking; Neither) | LAg (Positive: recent; Positive: long-term) | HIV-positive and unaware and antiretroviral not detectable or antiretroviral testing results missing |

| | | | | | | |
|---|---|---|---|---|---|---|
| | | | Partner, not live-in; Ex-spouse/Ex-partner; Friend/ Acquaintance; Sex worker; Sex worker client; Stranger; Other) | | | |
| Zimbabwe 2015 | In total, with how many different people have you had sex with in the last 12 months? | In the last 12 months, have you sold sex for money? (Yes; No)<br><br>In the last 12 months, have you paid money for sex? (Yes; No) | The last time you had sex with [initials], was a condom used? (Yes; No)<br><br>What is your relationship with your [initials] partner? (Husband/Wife; Live-in partner; Partner, not live-in; Ex-spouse/Ex-partner; Friend/ Acquaintance; Sex worker; Sex worker client; Stranger; Other) | The last time you had sex with [initials], did either of you drink alcohol beforehand? (Only I was drinking; Only partner was drinking; Both were drinking; Neither) | LAg (Positive: recent; Positive: long-term) | HIV-positive and unaware and antiretroviral not detectable or antiretroviral testing results missing |
| Zimbabwe 2020 | How many different people have you had sex with in the last 12 months? | - | The last time you had sex with [initials], was a condom used? (Yes; No)<br><br>What is your relationship with your [initials] partner? (Husband/Wife; Live-in partner; Partner, not live-in; Ex-spouse/Ex- | The last time you had sex with [initials], did either of you drink alcohol beforehand? (Only I was drinking; Only partner was drinking; Both were drinking; Neither) | LAg (Positive: recent; Positive: long-term) | HIV-positive and unaware and antiretroviral not detectable or antiretroviral testing results missing |

| | | | |
|---|---|---|---|
| | | | partner; Friend/ Acquaintance; Sex worker; Sex worker client; Stranger; Other) |

**Table S2.** Survey-weighted Prevalence and 95% Confidence Intervals of Patterns of Hazardous Alcohol Use across by Gender and Country, N=245,253.

| | Never drinker | Low risk-alcohol use | Hazardous alcohol use without binge | Binge alcohol use |
|---|---|---|---|---|
| | | Total | | |
| Eastern Africa | 75.0% (74.8%-75.2%) | 11.8% (11.7%-12.0%) | 5.4% (5.2%-5.5%) | 7.8% (7.7%-8.0%) |
| Southern Africa | 60.1% (59.6%-60.6%) | 16.4% (16.1%-16.8%) | 9.5% (9.2%-9.8%) | 14.0% (13.6%-14.3%) |
| | | Men | | |
| Eastern Africa | 61.4% (61.0-61.8%) | 17.8% (17.5%-18.1%) | 7.2% (7.0%-7.4%) | 13.5% (13.3%-13.8%) |
| Malawi | 63.0% (62.1%-63.8%) | 16.6% (15.9%-17.3%) | 6.1% (5.7%-6.5%) | 14.3% (13.7%-14.9%) |
| Mozambique | 57.1% (55.6%-58.7%) | 16.9% (15.8%-18.1%) | 8.2% (7.4%-9.1%) | 17.7% (16.5%-18.9%) |
| Tanzania | 67.2% (66.5%-68.0%) | 15.5% (15.0%-16.1%) | 8.2% (7.8%-8.6%) | 9.1% (8.6%-9.5%) |
| Uganda | 58.8% (57.7%-59.9%) | 23.3% (22.3%-24.3%) | 7.4% (6.8%-8.0%) | 10.5% (9.8%-11.2%) |
| Zambia | 59.4% (58.4%-60.5%) | 16.0% (15.2%-16.7%) | 6.8% (6.3%-7.4%) | 17.8% (17.0%-18.5%) |
| Zimbabwe | 55.3% (54.3%-56.3%) | 21.3% (20.4%-22.1%) | 6.7% (6.2%-7.2%) | 16.7% (15.9%-17.5%) |
| Southern Africa | 46.0% (45.3%-46.7%) | 21.6% (21.0%-22.2%) | 10.9% (10.4%-11.4%) | 21.5% (20.9%-22.1%) |
| Botswana | 48.2% (46.0%-50.2%) | 15.3% (13.7%-16.9%) | 6.3% (5.3%-7.3%) | 30.2% (28.1%-32.2%) |
| Eswatini | 60.6% (59.2%-62.0%) | 20.9% (19.8%-22.1%) | 4.2% (3.6%-4.8%) | 14.2% (13.2%-15.2%) |
| Lesotho | 35.3% (33.8%-36.7%) | 33.2% (31.8%-34.7%) | 15.0% (13.9%-15.4%) | 16.5% (15.4%-17.6%) |
| Namibia | 32.2% (30.6%-33.7%) | 25.2% (23.8%-26.6%) | 15.7% (14.4%-16.9%) | 27.0% (25.4%-28.5%) |
| South Africa | 48.7% (47.4%-50.2%) | 17.6% (16.5%-18.6%) | 12.2% (11.3%-13.2%) | 21.4% (20.2%-22.6%) |
| | | Women | | |
| Eastern Africa | 87.5% (87.3%-87.7%) | 6.4% (6.2%-6.5%) | 3.6% (3.4%-3.7%) | 2.6% (2.5%-2.7%) |
| Malawi | 95.4% (95.0%-95.7%) | 2.7% (2.4%-2.9%) | 0.8% (0.7%-1.0%) | 1.1% (0.9%-1.3%) |
| Mozambique | 80.0% (78.9%-81.1%) | 8.2% (7.4%-9.1%) | 6.4% (5.7%-7.0%) | 5.7% (5.1%-6.2%) |
| Tanzania | 84.3% (83.8%-84.8%) | 6.8% (6.4%-7.1%) | 6.2% (5.9%-6.5%) | 2.7% (2.5%-3.0%) |
| Uganda | 80.8% (80.1%-81.5%) | 11.4% (10.8%-11.9%) | 4.4% (4.0%-4.8%) | 3.4% (3.1%-3.8%) |
| Zambia | 86.2% (85.6%-86.8%) | 7.5% (7.0%-7.9%) | 2.8% (2.5%-3.1%) | 3.5% (3.2%-3.8%) |
| Zimbabwe | 93.3% (92.9%-93.7%) | 4.4% (4.1%-4.7%) | 1.1% (0.9%-1.2%) | 1.2% (1.1%-1.4%) |
| Southern Africa | 73.4% (72.9%-74.0%) | 11.5% (11.1%-11.9%) | 8.2% (7.8%-8.5%) | 6.9% (6.5%-7.2%) |
| Botswana | 72.4% (70.7%-74.2%) | 9.2% (7.9%-10.4%) | 6.3% (5.4%-7.3%) | 12.0% (10.8%-13.2%) |
| Eswatini | 88.3% (87.4%-89.1%) | 6.8% (6.1%-7.4%) | 2.3% (1.9%-2.6%) | 2.7% (2.3%-3.1%) |
| Lesotho | 66.3% (65.0%-67.5%) | 23.0% (21.9%-24.1%) | 7.9% (7.2%-8.6%) | 2.9% (2.4%-3.3%) |
| Namibia | 59.7% (58.3%-61.2%) | 13.2% (12.2%-14.2%) | 15.8% (14.6%-16.8%) | 11.4% (10.4%-12.3%) |
| South Africa | 75.8% (74.7%-76.9%) | 9.4% (8.7%-10.2%) | 8.3% (7.7%-9.0%) | 6.4% (5.8%-7.1%) |

**Table S3.** Unadjusted and Adjusted Odds Ratios, and 95% Confidence Intervals for the Association Between Patterns of Hazardous Alcohol Use and Risky Sexual Behaviors among People who are HIV Negative by Gender, n=245,253

| | 2+ partners in the past year[1] | | Sold or bought sex in the past year[1] | | Unprotected sex with a non-marital/non-cohabitating partner in past year[2] | | Alcohol use by self or partner at last sex with most recent partner[1] | |
|---|---|---|---|---|---|---|---|---|
| | Men | | | | | | | |
| | Unadj. OR (95%CI) | Adj OR (95% CI) | Unadj. OR (95%CI) | Adj. OR (95% CI) | Unadj. OR (95%CI) | Adj. OR (95% CI) | Unadj. OR (95%CI) | Adj. OR (95% CI) |
| Alcohol use | | | | | | | | |
| Non-drinker | Ref. | Ref. | Ref. | Ref. | Ref. | Ref. | Ref. | Ref. |
| Low-risk drinker | 1.35 (1.29-1.41) | 1.43 (1.37-1.50) | 1.38 (1.21-1.58) | 1.42 (1.23-1.65) | 0.91 (0.86-0.97) | 1.04 (0.98-1.11) | 12.95 (11.68-14.36) | 13.38 (12.05-14.87) |
| Hazardous non-binge drinker | 1.61 (1.51-1.71) | 1.81 (1.70-1.93) | 1.80 (1.54-2.10) | 1.82 (1.52-2.17) | 0.95 (0.88-1.03) | 1.25 (1.15-1.36) | 26.12 (23.40-29.16) | 28.19 (25.18-31.57) |
| Binge drinker | 1.76 (1.67-1.84) | 2.08 (1.98-2.19) | 2.64 (2.31-3.03) | 2.34 (1.98-2.75) | 1.12 (1.05-1.18) | 1.50 (1.41-1.60) | 33.16 (29.96-35.70) | 39.55 (35.54-44.01) |
| Age group | | | | | | | | |
| 15-24 years | 1.19 (1.14-1.24) | 1.09 (1.04-1.15) | 1.16 (1.03-1.31) | 1.12 (0.95-1.31) | 2.49 (2.37-2.62) | 2.56 (2.43-2.70) | 0.56 (0.52-0.61) | 0.71 (0.64-0.79) |
| 25-34 years | Ref. | Ref. | Ref. | Ref. | Ref. | Ref. | Ref. | Ref. |
| 35-49 years | 0.74 (0.71-0.78) | 0.80 (0.76-0.84) | 0.63 (0.55-0.72) | 0.66 (0.57-0.77) | 0.53 (0.50-0.57) | 0.51 (0.48-0.55) | 1.21 (1.14-1.30) | 1.26 (1.17-1.36) |
| 50+ years | 0.43 (0.40-0.45) | 0.49 (0.46-0.52) | 0.24 (0.20-0.30) | 0.33 (0.27-0.41) | 0.36 (0.34-0.40) | 0.34 (0.31-0.37) | 1.26 (1.18-1.36) | 1.41 (1.30-1.54) |
| Education | | | | | | | | |
| Primary or no education | Ref. | Ref. | Ref. | Ref. | Ref. | Ref. | Ref. | Ref. |
| Secondary | 1.02 (0.98-1.06) | 1.05 (1.01-1.09) | 0.87 (0.78-0.97) | 0.69 (0.60-0.78) | 1.04 (0.99-1.09) | 0.90 (0.86-0.95) | 0.82 (0.77-0.86) | 0.85 (0.80-0.91) |
| More than secondary | 0.93 (0.87-0.99) | 0.98 (0.91-1.05) | 0.57 (0.46-0.70) | 0.58 (0.45-0.74) | 0.93 (0.86-1.00) | 1.03 (0.94-1.12) | 0.77 (0.69-0.85) | 0.62 (0.55-0.70) |
| Employment Status | | | | | | | | |
| Unemployed or unable to work | Ref. | Ref. | Ref. | Ref. | Ref. | Ref. | Ref. | Ref. |
| Employed or student | 1.35 (1.30-1.40) | 1.32 (1.27-1.37) | 1.38 (1.24-1.53) | 1.45 (1.29-1.63) | 0.92 (0.88-0.96) | 0.94 (0.90-0.99) | 1.22 (1.16-1.29) | 1.03 (0.97-1.09) |
| Geographic area type | | | | | | | | |
| Urban | 0.99 (0.95-1.02) | 0.94 (0.90-0.98) | 1.09 (0.98-1.21) | 0.89 (0.79-1.02) | 1.12 (1.07-1.1) | 1.09 (1.04-1.15) | 1.09 (1.03-1.15) | 1.07 (1.00-1.15) |
| Rural | Ref. | Ref. | Ref. | Ref. | Ref. | Ref. | Ref. | Ref. |
| Ever married | | | | | | | | |
| Yes | Ref. | Ref. | Ref. | Ref. | | | Ref. | Ref. |
| No | 1.53 | 1.35 | 1.88 | 1.65 | | | 0.71 | 1.11 |

| | | | | | | | | |
|---|---|---|---|---|---|---|---|---|
| | (1.47-1.58) | (1.29-1.42) | (1.69-2.08) | (141-1.93) | | | (0.67-0.76) | (1.05-1.25) |
| Country | | | | | | | | |
| Eastern Africa | | | | | | | | |
| Malawi | 1.01 (0.90-1.14) | 1.08 (0.95-1.21) | 32.05 (23.37-43.96) | 30.79 (21.66-43.76) | 0.88 (0.76-1.01) | 0.75 (0.64-0.88) | 1.02 (0.85-1.22) | 1.56 (1.28-1.90) |
| Mozambique | 1.17 (1.02-1.34) | 1.22 (1.06-1.40) | - | - | 1.61 (1.38-1.88) | 1.34 (1.13-1.60) | 0.93 (0.75-1.14) | 1.15 (0.92-1.44) |
| Tanzania | 1.47 (1.31-1.65) | 1.77 (1.57-1.99) | 5.10 (3.81-6.82) | 5.25 (3.78-7.30) | 1.92 (1.67-2.20) | 2.04 (1.75-2.37) | 1.34 (1.12-1.60) | 2.52 (2.08-3.06) |
| Uganda | 1.49 (1.32-1.68) | 1.58 (1.40-1.79) | - | - | 1.69 (1.46-1.95) | 1.59 (1.35-1.86) | 2.33 (1.95-2.78) | 4.61 (3.78-5.62) |
| Zambia | 0.86 (0.76-0.97) | 0.83 (0.73-0.94) | 22.74 (16.60-31.14) | 29.84 (14.01-28.10) | 1.73 (1.50-1.99) | 1.52 (1.30-1.77) | 1.57 (1.31-1.88) | 2.34 (1.92-2.86) |
| Zimbabwe | 0.89 (0.79-1.01) | 0.99 (0.87-1.12) | 2.89 (2.12-3.95) | 3.35 (2.39-4.69) | 0.87 (0.75-1.00) | 0.88 (0.74-1.03) | 1.33 (1.11-1.59) | 1.93 (1.58-2.34) |
| Southern Africa | | | | | | | | |
| Botswana | Ref. | Ref. | - | - | Ref. | Ref. | Ref. | Ref. |
| Eswatini | 1.07 (0.94-1.22) | 1.18 (1.04-1.35) | Ref. | Ref. | 1.24 (1.06-1.44) | 1.17 (0.99-1.39) | 1.18 (0.95-1.48) | 2.12 (1.67-2.70) |
| Lesotho | 1.58 (1.39-1.80) | 1.54 (1.35-1.75) | 31.75 (22.12-45.55) | 25.18 (16.85-37.63) | 1.12 (0.96-1.31) | 0.94 (0.79-1.12) | 1.32 (1.09-1.61) | 1.28 (1.04-1.58) |
| Namibia | 1.07 (0.93-1.23) | 0.86 (0.75-0.99) | 23.36 (14.82-36.82) | 18.72 (11.44-30.63) | 1.49 (1.27-1.75) | 1.24 (1.04-1.48) | - | - |
| South Africa | 0.65 (0.56-0.74) | 0.62 (0.54-0.72) | 24.44 (15.80-37.82) | 23.88 (15.00-38.03) | 0.99 (0.82-1.18) | 0.72 (0.59-0.89) | 1.28 (1.04-1.56) | 1.30 (1.05-1.62) |
| Age at first sex | | | | | | | | |
| ≤15 years | 1.74 (1.68-1.81) | 1.52 (1.45-1.58) | 2.29 (2.06-2.55) | 1.37 (1.22-1.55) | 1.84 (1.76-1.93) | 1.35 (1.29-1.42) | 1.16 (1.10-1.23) | 1.18 (1.10-1.26) |
| >15 years | Ref. | Ref. | Ref. | Ref. | Ref. | Ref. | Ref. | Ref. |
| | | | | Women | | | | |
| | Unadjusted OR (95%CI) | Adjusted OR (95% CI) | Unadjusted OR (95%CI) | Adjusted OR (95% CI) | Unadjusted OR (95%CI) | Adjusted OR (95% CI) | Unadjusted OR (95%CI) | Adjusted OR (95% CI) |
| Alcohol use | | | | | | | | |
| Non-drinker | Ref. | Ref. | Ref. | Ref. | Ref. | Ref. | Ref. | Ref. |
| Low-risk drinker | 2.58 (2.35-2.82) | 2.38 (2.15-2.62) | 1.64 (1.26-2.13) | 1.65 (1.23-2.21) | 1.56 (1.45-1.67) | 1.49 (1.38-1.60) | 2.61 (2.44-2.80) | 3.02 (2.81-3.25) |
| Hazardous non-binge drinker | 3.44 (3.10-3.82) | 3.57 (3.18-4.00) | 2.34 (1.80-3.04) | 2.17 (1.62-2.91) | 1.82 (1.67-1.98) | 1.73 (1.57-1.89) | 3.91 (3.60-4.24) | 5.25 (4.80-5.73) |
| Binge drinker | 4.35 (3.90-4.87) | 4.36 (3.86-4.93) | 5.58 (4.07-7.66) | 4.45 (2.82-7.04) | 2.05 (1.87-2.24) | 1.92 (1.75-2.12) | 4.55 (4.16-5.00) | 6.90 (6.23-7.64) |
| Age group | | | | | | | | |
| 15-24 years | 1.44 | 1.27 | 1.84 | 1.24 | 1.93 | 2.03 | 0.67 | 0.74 |

| | | | | | | | | |
|---|---|---|---|---|---|---|---|---|
| | (1.34-1.54) | (1.17-1.37) | (1.54-2.20) | (1.01-1.52) | (1.84-2.02) | (1.93-2.13) | (0.64-0.72) | (0.70-0.80) |
| 25-34 years | Ref. | Ref. | Ref. | Ref. | Ref. | Ref. | Ref. | Ref. |
| 35-49 years | 0.58<br>(0.52-0.63) | 0.61<br>(0.56-0.68) | 0.80<br>(0.65-1.00) | 0.73<br>(0.58-0.92) | 0.81<br>(0.76-0.86) | 0.83<br>(0.78-0.88) | 1.23<br>(1.17-1.30) | 1.16<br>(1.10-1.23) |
| 50+ years | 0.09<br>(0.07-0.11) | 0.10<br>(0.08-0.12) | 0.18<br>(0.12-0.28) | 0.17<br>(0.11-0.27) | 0.42<br>(0.38-0.46) | 0.47<br>(0.43-0.52) | 1.31<br>(1.22-1.40) | 1.19<br>(1.11-1.29) |
| Education | | | | | | | | |
| No education and Primary | Ref. | Ref. | Ref. | Ref. | Ref. | Ref. | Ref. | Ref. |
| Secondary | 1.53<br>(1.43-1.64) | 1.00<br>(0.92-1.09) | 0.70<br>(0.59-0.83) | 0.61<br>(0.49-0.76) | 1.47<br>(1.41-1.53) | 1.30<br>(1.24-1.37) | 0.64<br>(0.61-0.67) | 0.76<br>(0.72-0.81) |
| More than secondary | 1.72<br>(1.54-1.93) | 0.89<br>(0.78-1.01) | 0.17<br>(0.10-0.28) | 0.15<br>(0.07-0.31) | 1.38<br>(1.28-1.49) | 1.19<br>(1.08-1.30) | 0.61<br>(0.55-0.68) | 0.59<br>(0.52-0.66) |
| Employment Status | | | | | | | | |
| Unemployed or unable to work | Ref. | Ref. | Ref. | Ref. | Ref. | Ref. | Ref. | Ref. |
| Employed or student | 1.53<br>(1.43-1.63) | 1.40<br>(1.30-1.49) | 1.52<br>(1.31-1.77) | 1.65<br>(1.39-1.96) | 1.29<br>(1.24-1.34) | 1.30<br>(1.24-1.36) | 1.25<br>(1.20-1.31) | 1.17<br>(1.11-1.22) |
| Geographic area type | | | | | | | | |
| Urban | 1.70<br>(1.60-1.81) | 1.31<br>(1.22-1.41) | 1.21<br>(1.03-1.41) | 0.94<br>(0.78-1.13) | 1.52<br>(1.46-1.58) | 1.28<br>(1.22-1.34) | 0.86<br>(0.82-0.90) | 0.95<br>(0.90-1.00) |
| Rural | Ref. | Ref. | Ref. | Ref. | Ref. | Ref. | Ref. | Ref. |
| Ever married | | | | | | | | |
| Yes | Ref. | Ref. | Ref. | Ref. | | | Ref. | Ref. |
| No | 2.97<br>(2.78-3.17) | 2.09<br>(1.93-2.27) | 3.64<br>(3.12-4.25) | 3.07<br>(2.50-3.76) | | | 0.55<br>(0.51-0.59) | 0.82<br>(0.76-0.89) |
| Country | | | | | | | | |
| Eastern Africa | | | | | | | | |
| Malawi | 0.33<br>(0.28-0.39) | 0.68<br>(0.56-0.82) | 17.04<br>(12.82-22.65) | 14.46<br>(10.52-19.88) | 0.55<br>(0.49-0.62) | 0.75<br>(0.65-0.85) | 1.51<br>(1.26-1.81) | 1.99<br>(1.63-2.41) |
| Mozambique | 0.44<br>(0.36-0.55) | 0.62<br>(0.50-0.77) | - | - | 1.07<br>(0.93-1.22) | 1.19<br>(1.03-1.37) | 1.03<br>(0.83-1.28) | 0.94<br>(0.75-1.19) |
| Tanzania | 0.70<br>(0.60-0.81) | 1.25<br>(1.06-1.48) | Ref. | Ref. | 1.11<br>(0.99-1.25) | 1.41<br>(1.25-1.60) | 2.01<br>(1.68-2.39) | 2.03<br>(1.68-2.45) |
| Uganda | 0.54<br>(0.45-0.63) | 0.81<br>(0.67-0.98) | - | - | 1.09<br>(0.99-1.25) | 1.22<br>(1.07-1.39) | 4.00<br>(3.35-4.78) | 4.27<br>(3.52-5.17) |
| Zambia | 0.31<br>(0.26-0.37) | 0.43<br>(0.35-0.52) | 6.72<br>(4.62-9.77) | 4.25<br>(2.86-6.31) | 0.91<br>(0.81-1.03) | 1.06<br>(0.93-1.20) | 3.06<br>(2.57-3.66) | 3.75<br>(3.09-4.53) |
| Zimbabwe | 0.29<br>(0.24-0.34) | 0.66<br>(0.55-0.80) | 0.13<br>(0.09-0.18) | 0.20<br>(0.14-0.28) | 0.44<br>(0.39-0.50) | 0.54<br>(0.47-0.61) | 2.26<br>(1.90-2.70) | 3.35<br>(2.77-4.05) |
| Southern Africa | | | | | | | | |
| Botswana | Ref. | Ref. | - | - | Ref. | Ref. | Ref. | Ref. |

| | | | | | | | | |
|---|---|---|---|---|---|---|---|---|
| Eswatini | 0.62<br>(0.52-0.74) | 1.04<br>(0.85-1.26) | 0.06<br>(0.03-0.10) | 0.07<br>(0.03-0.13) | 1.10<br>(0.96-1.25) | 1.29<br>(1.13-1.48) | 1.70<br>(1.37-2.10) | 2.47<br>(1.97-3.10) |
| Lesotho | 0.92<br>(0.78-1.09) | 1.35<br>(1.12-1.62) | 10.67<br>(6.49-17.53) | 10.25<br>(5.61-18.74) | 0.58<br>(0.50-0.67) | 0.57<br>(0.49-0.66) | 1.63<br>(1.34-1.99) | 1.52<br>(1.23-1.87) |
| Namibia | 0.66<br>(0.55-0.81) | 0.49<br>(0.40-0.60) | 16.93<br>(7.90-36.28) | 7.55<br>(3.42-16.65) | 1.35<br>(1.19-1.55) | 1.19<br>(1.04-1.37) | - | - |
| South Africa | 0.35<br>(0.29-0.42) | 0.38<br>(0.31-0.47) | 7.99<br>(5.36-11.91) | 6.76<br>(4.06-11.26) | 1.12<br>(0.95-1.31) | 0.93<br>(0.79-1.11) | 0.74<br>(0.59-0.92) | 0.73<br>(0.58-0.92) |
| Age at first sex | | | | | | | | |
| ≤15 years | 1.71<br>(1.59-1.83) | 1.83<br>(1.70-1.98) | 2.45<br>(2.09-2.86) | 1.43<br>(1.19-1.73) | 1.27<br>(1.21-1.33) | 1.28<br>(1.21-1.34) | 1.40<br>(1.33-1.48) | 1.19<br>(1.12-1.26) |
| >15 years | Ref. | Ref. | Ref. | Ref. | Ref. | Ref. | Ref. | Ref. |

Model 1: Adjusted for age group, education, employment status, geographic area type, ever married, country, and age at first sex
Model 2: Adjusted for age group, education, employment status, geographic area type, country, and age at first sex
Note: Gray boxes indicate that a variable was not included in the model; Dash indicates that variable was not included in the country-specific survey

**Table S4.** Unadjusted and Adjusted Odds Ratios and 95% Confidence Intervals for the Association Between Patterns of Hazardous Alcohol Use and Recent HIV Infection by Gender, n=251,931

| | Recent HIV Infection | | | | Undiagnosed HIV Infection | | | |
|---|---|---|---|---|---|---|---|---|
| | Men | | Women | | Men | | Women | |
| | Unadjusted OR (95%CI) | Adjusted OR (95% CI) | Unadjusted OR (95%CI) | Adjusted OR (95% CI) | Unadjusted OR (95%CI) | Adjusted OR (95% CI) | Unadjusted OR (95%CI) | Adjusted OR (95% CI) |
| Alcohol use | | | | | | | | |
| Non-drinker | Ref. | Ref. | Ref. | Ref. | Ref. | Ref. | Ref. | Ref. |
| Low-risk drinker | 1.06 (0.61-1.83) | 0.95 (0.54-1.67) | 1.44 (0.95-2.18) | 1.28 (0.82-1.99) | 1.45 (1.28-1.64) | 1.32 (1.16-1.50) | 1.34 (1.19-1.52) | 1.28 (1.13-1.46) |
| Hazardous non-binge drinker | 1.42 (0.71-2.86) | 1.24 (0.60-2.56) | 1.51 (0.87-2.62) | 1.63 (0.90-2.94) | 1.62 (1.38-1.90) | 1.41 (1.19-1.65) | 1.50 (1.30-1.73) | 1.38 (1.19-1.61) |
| Binge drinker | 1.77 (1.03-3.05) | 1.61 (0.92-2.81) | 1.60 (0.88-2.89) | 1.63 (0.89-2.98) | 1.87 (1.66-2.12) | 1.52 (1.34-1.72) | 1.79 (1.53-2.09) | 1.55 (1.31-1.82) |
| Age group | | | | | | | | |
| 15-24 years | 0.42 (0.23-0.77) | 3.50 (1.73-7.09) | 1.12 (0.84-1.48) | 1.06 (0.77-1.45) | 0.25 (0.21-0.30) | 0.29 (0.24-0.35) | 0.61 (0.55-0.67) | 0.54 (0.49-0.60) |
| 25-34 years | Ref. | Ref. | Ref. | Ref. | Ref. | Ref. | Ref. | Ref. |
| 35-49 years | 0.97 (0.58-1.64) | 1.21 (0.69-2.12) | 0.65 (0.46-0.93) | 0.66 (0.45-0.95) | 1.34 (1.19-1.49) | 1.28 (1.14-1.45) | 0.99 (0.91-1.09) | 1.00 (0.91-1.10) |
| 50+ years | 0.77 (0.42-1.40) | 0.93 (0.45-1.92) | 0.31 (0.17-0.54) | 0.31 (0.17-0.56) | 0.66 (0.57-0.75) | 0.59 (0.50-0.69) | 0.54 (0.49-0.61) | 0.51 (0.45-0.58) |
| Education | | | | | | | | |
| No education and Primary | Ref. | Ref. | Ref. | Ref. | Ref. | Ref. | Ref. | Ref. |
| Secondary | 1.21 (0.78-1.86) | 1.08 (0.67-1.74) | 1.82 (1.41-2.36) | 1.08 (0.76-1.52) | 1.09 (0.99-1.20) | 0.84 (0.75-0.94) | 1.38 (1.28-1.49) | 0.85 (0.77-0.963) |
| More than secondary | 1.19 (0.56-2.54) | 0.69 (0.29-1.67) | 1.08 (0.63-1.84) | 0.45 (0.24-0.86) | 0.80 (0.66-0.97) | 0.49 (0.39-0.60) | 0.97 (0.83-1.14) | 0.50 (0.42-0.60) |
| Employment Status | | | | | | | | |
| Unemployed or unable to work | Ref. | Ref. | Ref. | Ref. | Ref. | Ref. | Ref. | Ref. |
| Employed or student | 1.13 (0.74-1.72) | 1.00 (0.64-1.57) | 1.26 (0.97-1.63) | 1.32 (0.99-1.74) | 1.22 (1.11-1.34) | 1.09 (0.98-1.20) | 1.13 (1.05-1.22) | 1.09 (1.01-1.18) |
| Geographic area type | | | | | | | | |
| Urban | 1.36 (0.89-2.07) | 1.37 (0.86-2.19) | 1.39 (1.08-1.80) | 1.30 (0.96-1.78) | 1.27 (1.16-1.40) | 1.23 (1.10-1.37) | 1.45 (1.35-1.56) | 1.28 (1.18-1.39) |
| Rural | Ref. | Ref. | Ref. | Ref. | Ref. | Ref. | Ref. | Ref. |
| Ever married | | | | | | | | |
| Yes | Ref. | Ref. | Ref. | Ref. | Ref. | Ref. | Ref. | Ref. |
| No | 0.98 | 1.54 | 1.71 | 1.14 | 0.60 | 0.82 | 1.57 | 1.51 |

| | | | | | | | | |
|---|---|---|---|---|---|---|---|---|
| | (0.62-1.54) | (0.82-2.89) | (1.30-2.24) | (0.82-1.58) | (0.53-0.67) | (0.71-0.95) | (1.45-1.70) | (1.36-1.67) |
| Country | | | | | | | | |
| Eastern Africa | | | | | | | | |
| Malawi | 1.13 | 1.21 | 1.18 | 1.27 | 1.31 | 1.34 | 1.47 | 1.76 |
| | (0.52-2.46) | (0.54-2.70) | (0.72-1.4) | (0.76-2.12) | (0.90-1.91) | (0.91-1.96) | (1.03-2.09) | (1.23-2.53) |
| Mozambique | 1.40 | 1.37 | 1.89 | 1.78 | 2.48 | 2.52 | 3.52 | 3.89 |
| | (0.50-3.91) | (0.49-3.79) | (1.01-3.56) | (0.94-3.36) | (1.67-3.69) | (1.69-3.76) | (2.47-5.03) | (2.69-5.56) |
| Tanzania | Ref. | Ref. | Ref. | Ref. | 0.86 | 0.79 | 1.21 | 1.28 |
| | | | | | (0.59-1.25) | (0.54-1.17) | (0.85-1.71) | (0.89-1.83) |
| Uganda | 1.65 | 1.79 | 1.49 | 1.40 | 0.84 | 0.85 | 1.02 | 1.13 |
| | (0.69-3.93) | (0.75-4.31) | (0.86-2.58) | (0.81-2.42) | (0.56-1.27) | (0.56-1.29) | (0.70-1.48) | (0.77-1.66) |
| Zambia | 1.12 | 1.03 | 2.42 | 2.26 | 1.63 | 1.68 | 2.58 | 2.79 |
| | (0.53-2.38) | (0.47-2.25) | (1.55-3.79) | (1.44-3.56) | (1.12-2.37) | (1.0915-2.45) | (1.82-3.64) | (1.96-3.98) |
| Zimbabwe | 1.85 | 1.85 | 1.72 | 1.92 | 2.20 | 2.17 | 2.38 | 3.03 |
| | (0.90-3.80) | (0.86-4.03) | (1.08-2.74) | (1.20-3.07) | (1.53-3.19) | (1.49-3.15) | (1.69-3.36) | (2.13-4.33) |
| Southern Africa | | | | | | | | |
| Botswana | 1.41 | 0.40 | 1.27 | 1.23 | Ref. | Ref. | Ref. | Ref. |
| | (0.85-2.36) | (0.20-0.77) | (0.49-3.29) | (0.43-3.53) | | | | |
| Eswatini | 3.60 | 3.25 | 4.24 | 5.24 | 2.87 | 3.41 | 2.74 | 3.56 |
| | (1.64-7.93) | (1.35-7.83) | (2.62-6.85) | (3.01-9.11) | (1.96-4.19) | (2.33-5.01) | (1.91-3.92) | (2.46-5.14) |
| Lesotho | 3.51 | 3.08 | 3.16 | 3.26 | 1.94 | 1.85 | 2.55 | 2.96 |
| | (1.64-7.71) | (1.34-7.04) | (1.89-5.28) | (1.90-5.60) | (1.31-2.89) | (1.23-2.76) | (1.78-3.68) | (2.04-4.29) |
| Namibia | 1.30 | 0.96 | 2.03 | 1.62 | 1.66 | 1.55 | 1.56 | 1.38 |
| | (0.44-3.84) | (0.30-3.07) | (1.10-3.74) | (0.83-3.13) | (1.10-2.50) | (1.02-2.33) | (1.06-2.28) | (0.94-2.04) |
| South Africa | - | - | - | - | 2.85 | 2.86 | 4.84 | 4.72 |
| | | | | | (1.94-4.16) | (1.95-4.18) | (3.42-6.83) | (3.33-6.69) |
| Age at first sex | | | | | | | | |
| ≤15 years | Ref. | Ref. | Ref. | Ref. | Ref. | Ref. | Ref. | Ref. |
| >15 years | 1.16 | 1.41 | 1.27 | 1.33 | 0.70 | 0.91 | 0.97 | 1.12 |
| | (0.72-1.86) | (0.86-2.32) | (0.94-1.70) | (0.97-1.82) | (0.62-0.79) | (0.81-1.03) | (0.89-1.06) | (1.02-1.23) |

Models adjusted for age group, education, employment status, geographic area type, ever married, and age at first sex
Note: Dash indicates that variable was not included in the country-specific survey
